# Classification of *GBA1* variants in Parkinson’s disease; the *GBA1*-PD browser

**DOI:** 10.1101/2022.09.27.22280421

**Authors:** Sitki Cem Parlar, Francis P. Grenn, Jonggeol Jeffrey Kim, Cornelis Blauwendraat, Ziv Gan-Or

## Abstract

**Background:** *GBA1* variants are among the most common genetic risk factors for Parkinson’s Disease (PD). *GBA1* variants can be classified into three categories based on their role in Gaucher’s Disease (GD) or PD: severe, mild, and risk variant (for PD).

**Objectives:** This paper aims to generate and share a comprehensive database for *GBA1* variants reported in PD to support future research and clinical trials.

**Methods:** We performed a literature search for all *GBA1* variants that have been reported in PD. The data has been standardized and complimented with variant classification, Odds Ratio (OR) if available and other data.

**Results:** We found 371 *GBA1* variants reported in PD: 22 mild, 84 severe, 3 risk variants, and 262 of unknown status. We created a browser, containing up-to-date information on these variants (https://pdgenetics.shinyapps.io/GBA1Browser/).

**Conclusions:** The classification and browser presented in this work should inform and support basic, translational, and clinical research on *GBA1*-PD.

## INTRODUCTION

The risk, onset, and progression of PD are influenced by a multitude of factors including aging, environmental exposures, and genetic background.^1^ The underlying pathobiological mechanisms influencing PD risk are not fully understood; however, multiple genetic loci for PD risk have been identified in the human genome, as well as genes involved in rare Mendelian forms of PD. Notable genes implicated in PD include: *SNCA, LRRK2, PRKN*, and *PINK1*, among others.^1^ One of the most important genes in PD is *GBA1*, as 5-20% of PD patients in different populations carry variants in this gene.^2^

*GBA1* encodes the lysosomal enzyme glucocerebrosidase (GCase), responsible for hydrolyzing glucosylceramide and glucosylsphingosine.^3^ Pathogenic biallelic *GBA1* variants cause a lysosomal storage disorder, Gaucher Disease (GD), which can be classified as type I (mild, non-neuronopathic form of GD), type II or type III (severe, neuronopathic forms of GD). Accordingly, *GBA1* variants can be classified as mild or severe, based on the type of GD that they lead to in a homozygous state.^2^

The association between *GBA1* variants and PD originates from clinical observations, reporting that some GD patients had been also displaying clinical signs of PD.^4-6^ Genetic studies subsequently showed that *GBA1* variants are common risk factors in PD in various populations^2, 7^ and that the type of *GBA1* variants, mild or severe, is associated with differential risk and progression of PD. Carriers of severe *GBA1* variants have higher risk for PD, earlier age at onset^2^, and their motor and cognitive decline is faster.^8, 9^ However, the majority of *GBA1* variant carriers do not develop PD, as the penetrance of heterozygous *GBA1* variants in most PD populations ranges between 10-30%.^10-12^ This is much higher compared to a 6.6% reported life-time risk for PD observed in the overall population PD,^13-16^ yet the mechanism by which *GBA1* variants cause or increase the susceptibly for PD is still unknown.^17^

Since the association was first established between *GBA1* variants and PD, there has been extensive research on genotype-phenotype correlations. The list of *GBA1* variants reported in PD cases has grown overtime, and is not fully overlapping that of GD. Notably, the p.E326K and p.T369M variants, which do not cause GD, are risk factors of PD.^18, 19^ Since more and more clinical trials targeting individuals with PD and *GBA1* variants are being performed and planned, it is important to gather data on *GBA1* variants to inform the design of these trials and other clinical and functional studies. For example, since carriers of severe *GBA1* variants are likely to progress faster, it will be important that they will be equally represented in the treatment and placebo arms of trials. Here, we compiled a list of all *GBA1* variants reported in individuals with PD to date. We generated an online browser to mine data on these variants, including the severity if known among other important information, and we will continue to update this resource.

## METHODS

### Literature Search Criteria

For the purposes of creating the *GBA1*-PD browser, we searched for all studies that reported *GBA1* variants in PD populations. An initial search on PubMed was done including the variations of the following keywords: “*GBA*,” “*GBA1*,” and “glucocerebrosidase,” and “Parkinson’s,” and “parkinsonism.” The search included papers published from the year 2004, when *GBA*1 was first suggested as a probable risk factor for PD onset,^20^ up to April 2022, when the literature search was conducted. As a result of this initial search, 1128 papers were found. After removing meta-analyses and review papers from the list, the remaining 834 papers were thoroughly screened. The screening then filtered for studies involving case-control or case-only PD populations with data on common and rare *GBA1* variants. The final list after this second screening step consisted of 86 papers in total (Figure 1).

**Figure 1.**
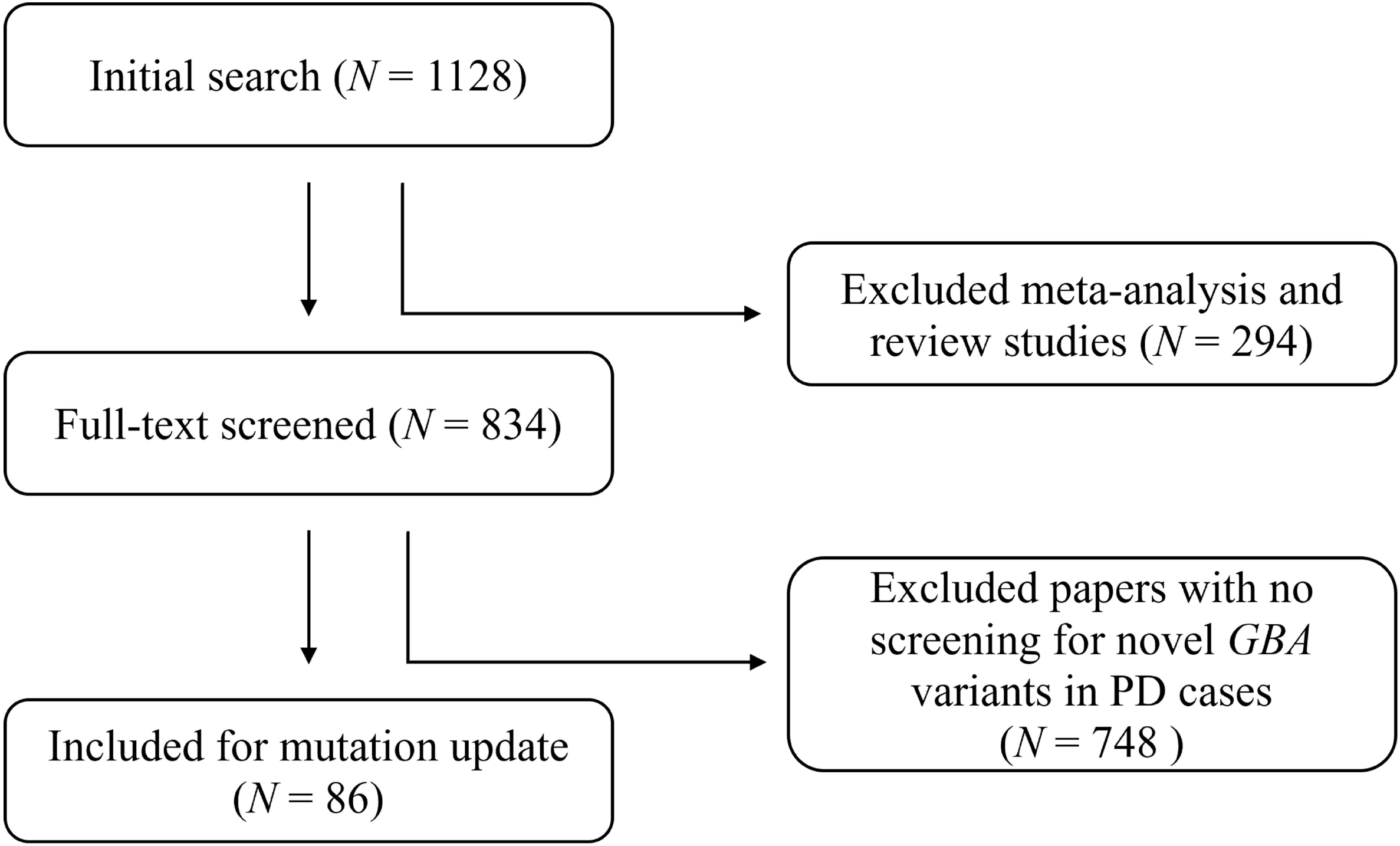
Flow chart of the literature search. Representation of the literature search in flowchart format, involving two sequential steps for screening: the removal of meta-analyses and review papers, and the removal of papers that do not contain novel *GBA1* variants in PD patients. N=number of studies

### Quality Control

*GBA1* variants collected from the final list of literature were then validated based on the sequencing data of the *GBA1* gene in ensemble.org. ^21^ In addition, variant information was revised according to the Human Genome Variation Society (HGVS) guidelines for variants nomenclature.^22^

### Clinical Classification of *GBA1* variants

As mentioned earlier, the classification of *GBA1* variants as mild or severe is based on GD: mild mutations cause GD type I and severe mutations cause GD type II or III. This classification is also important in PD, as carriers of severe *GBA1* variants have a higher risk of PD, earlier AAO,^2^ and faster cognitive and motor decline.^8, 9, 23^ There are also *GBA1* variants that do not cause GD but are associated with increased risk of PD, such as p.E326K and p.T369M.^18, 19^ We therefore performed a literature search for each variant to find if it was reported in GD and if its severity was interpretable. We then classified the variants accordingly as mild (causing GD type I), severe (causing GD type II or III), risk variant (variants that are associated with risk of PD but do not cause GD), and unknown.

### Construction of browser and data sharing

In order to make the information openly accessible, we built the *GBA1*-PD Browser (https://pdgenetics.shinyapps.io/GBA1Browser/). This browser was built using R Shiny and includes specific information pertaining to each variant reported in PD as follows: variant name, full length name, clinical classification (i.e. mild, severe, risk variant, or unknown), rsID, genome base pair position (hg19), exon number, allele frequency in gnomAD,^24^ CADD PHRED-scaled score,^25^ GERP scores,^26^ and the manuscript that reported the variant. The data was generated and compiled before being added into the browser.

## RESULTS

### Identification and classification of *GBA1* variants

From the 86 studies that reported *GBA1* variants in PD cases, we identified 371 unique variants reported to date (https://pdgenetics.shinyapps.io/GBA1Browser/). Of the 371 variants, 22 were identified as mild (type-I GD causing) variants, 84 were identified as severe (type-II and III GD causing) variants and 3 were identified as non-GD risk variants for PD. The risk variants found were biallelic and heterozygous forms of p.E326K, p.T369M, and p.E388K. The remaining 262 were classified as unknown due to lack of information on their GD pathology and/or PD risk associations. Figure 2 depicts the distribution of *GBA1* variants per exon, and the location of the most common variants associated with PD. Among the 86 studies included in this analysis, 16 reported variant specific odds ratio (OR) data for PD risk (Table 1).

**Table 1.**
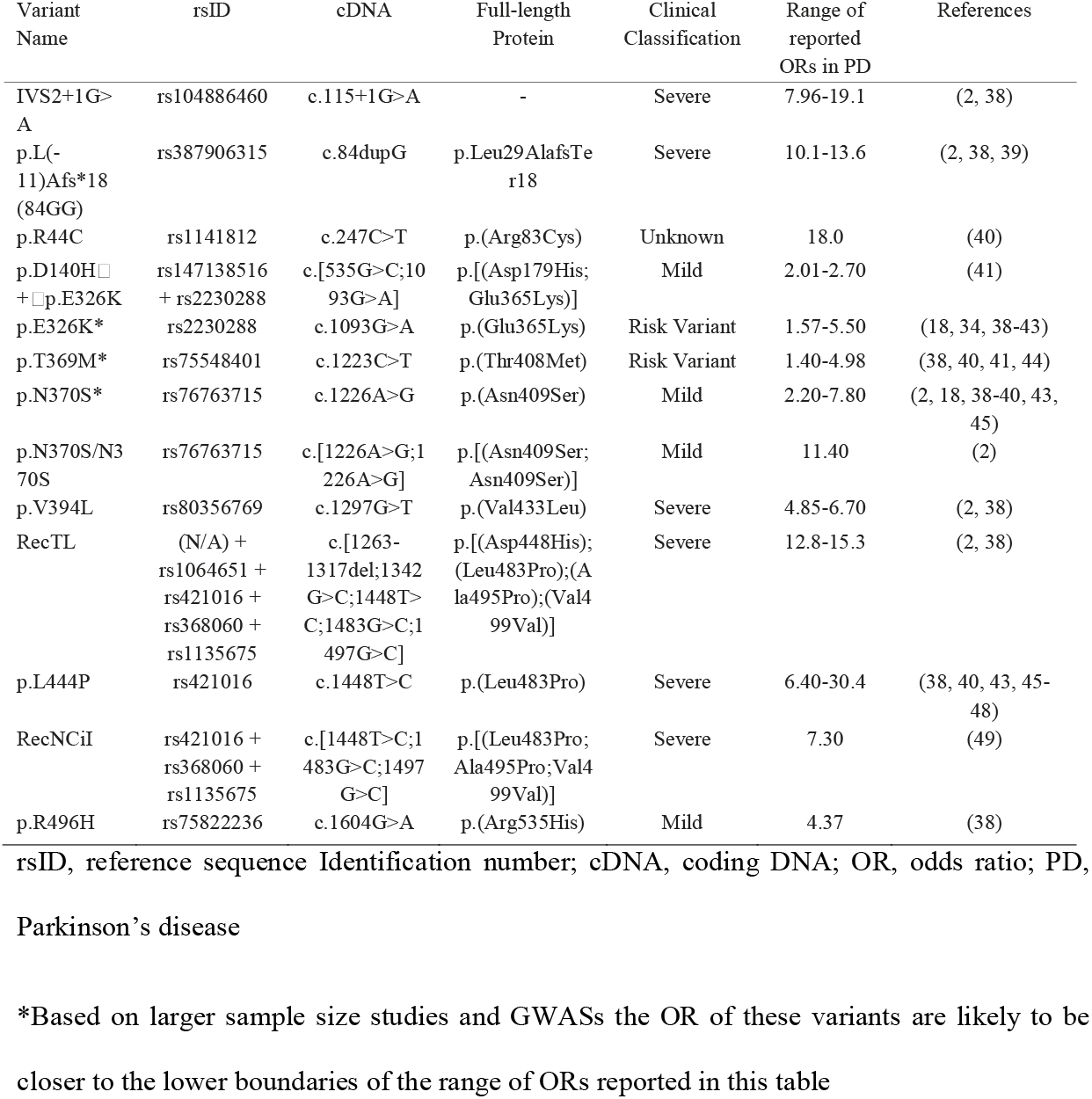
Odds Ratios of Parkinson’s Disease for specific *GBA1* variants.

**Figure 2.**
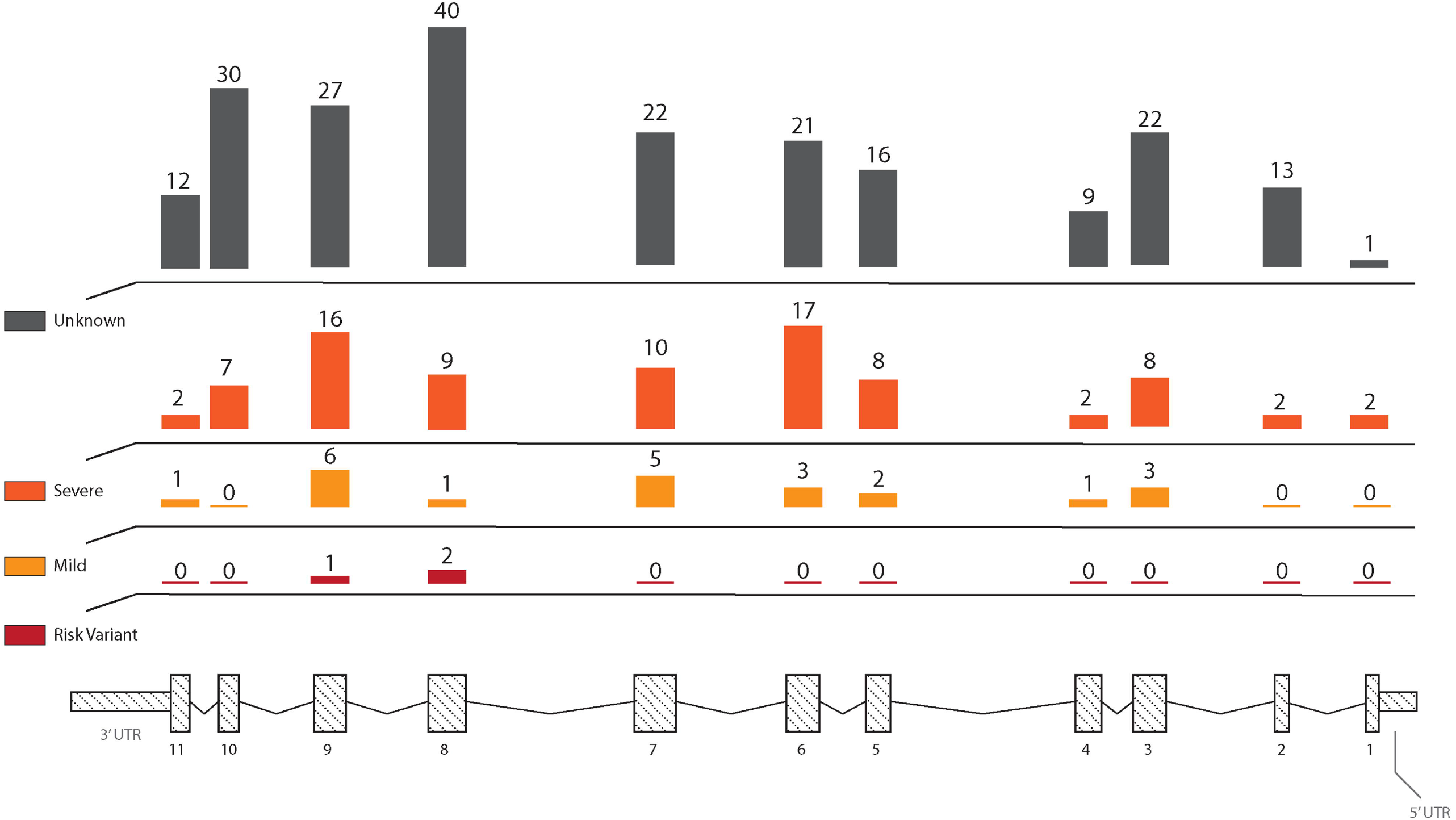
Distribution of *GBA1* variants reported in PD across the gene. Representation of the number of unique reported *GBA1* variants on each exon (numbered 1-11) by clinical classification (Unknown, Severe, Mild, and Risk Variant).

### The *GBA1*-PD browser

The *GBA1*-PD browser is a public-facing database created to assist researchers in finding *GBA1* variants relevant to PD risk. The previously described 371 variants can be searched and filtered through using hg19/hg38 position, protein consequence, rsID, clinical classification, and exon number. An interactive plot displays the location of the included variants in *GBA1*, and groups variants by their severity. Additional relevant information including reported allele frequency in gnomAD, CADD score, GERP score, and the reporting manuscript are available for each variant.

## DISCUSSION

The *GBA1* browser generated through this work could serve as a valuable resource for researchers, clinicians, and for design of clinical trials. *GBA1* variant carriers are at higher risk for PD with a penetrance of 10-30%,^10-12^ compared to the overall population who carry up to 6.6% lifetime risk for developing PD.^13-16^ The latter estimation includes carriers of *GBA1* variants, therefore the lifetime risk for non-carriers of *GBA1* variants is likely lower. There are notable differences across ancestries as to how common *GBA1* variant carriers are. For instance, the prevalence of *GBA1* variants in the Ashkenazi Jewish PD population is around 20%,^2, 27^ whereas for Chinese PD cases it is found at a much lower prevalence rate of around 5.4-8.4%.^28-30^ The frequencies of specific *GBA1* variants also differ across populations. The most frequent *GBA1* variants in Ashkenazi Jews with PD is p.N370S, while in European populations it is mostly p.E326K or p.T369M, and in Asian populations it is p.L444P.^2, 24^

These different variants also represent different categories of variants, classified based on their effect in GD. The p.N370S variant is a mild variant associated with GD type I,^31, 32^ p.E326K and p.T369M are risk variants for PD that do not cause GD,^19, 33, 34^ and p.L444P is a severe variant associated with the severe form of GD.^35-37^ While the odds ratio for PD associated with risk variants (i.e. p.E326K and p.T369M) is below 2,^19, 33^ the odds ratio of mild *GBA1* variants is above 2, and the odds ratio of severe *GBA* variants may reach above 10.^2^ More importantly, the variants may have different effects on PD progression, as motor symptoms and cognition seem to decline faster among carriers of severe *GBA1* variants.^8, 9, 23^ These observations could be especially important for clinical trials on *GBA1*-PD. If the treatment and placebo arms of a trial will not be balanced in terms of the composition of severe and mild variants in each arm, it is possible that one arm will progress faster than the other, which may lead to either false positive or negative results for a trial. Furthermore, trials that only include severe *GBA1* variant carriers may require a shorter trial duration and a smaller trial population, while trials that include mild or risk variant carriers may require a longer trial duration and a larger trial population. The *GBA1*-PD browser we generated could be helpful for designing such trials, and it will be kept up to date as more information becomes available on *GBA1* variants in GD and PD.

## Data Availability

All data collected for the study is publicly available through the GBA1-PD browser we generated at https://pdgenetics.shinyapps.io/GBA1Browser/

## ACKNOWLEDGMENTS

We thank those who have been a part of the referenced clinical studies that form the basis of this paper and its accompanying online database. This work has been supported by grants from the Michael J. Fox Foundation and the Canadian Consortium on Neurodegeneration in Aging (CCNA). This work was supported in part by the Intramural Research Programs of the National Institute on Aging (NIA).

## AUTHORS’ ROLES

S. Parlar: Execution of the research project. Literature search and review. Collection and organization of data. Making tables and figures, writing, and editing for the manuscript.

Z. Gan-Or: Conception of the research project and supervision in its execution. Review and critique of the manuscript.

C. Baluwendraat, F. P. Grenn, and J. J. Kim: Conception, organization, and execution of the online PD *GBA1* variant searching tool database. Review and critique of the manuscript.

## FINANCIAL DISCLOSURES OF ALL AUTHORS OF THE PRECEDING 12 MONTHS

S. C. P, F. P. G., J. J. K. and C. B. have nothing to report.

Z. G.-O. received consultancy fees from Lysosomal Therapeutics Inc. (LTI), Idorsia, Prevail Therapeutics, Inceptions Sciences (now Ventus), Neuron23, Handl Therapeutics, UCB, Ono Therapeutics, Denali, Bial Biotech, Guidepoint, Lighthouse, and Deerfield.

## REFERENCES

1. Blauwendraat C, Nalls MA, Singleton AB. The genetic architecture of Parkinson’s disease. Lancet Neurol 2020;19(2):170–178.

2. Gan-Or Z, Amshalom I, Kilarski LL, et al. Differential effects of severe vs mild GBA mutations on Parkinson disease. Neurology 2015;84(9):880–887.

3. Do J, McKinney C, Sharma P, Sidransky E. Glucocerebrosidase and its relevance to Parkinson disease. Mol Neurodegener 2019;14(1):36.

4. Philippart M, Rosenstein B, Menkes JH. Isolation and Characterization of the Main Splenic Glycolipids in the Normal Organ and in Gaucher’s Disease: Evidence for the Site of Metabolic Block. J Neuropathol Exp Neurol 1965;24:290–303.

5. Rabey JM, Vardi J, Askenazi JJ, Streifler M. L-tryptophan administration in L-dopa-induced hallucinations in elderly Parkinsonian patients. Gerontology 1977;23(6):438–444.

6. Roy M, Boyer L, Barbeau A. A prospective study of 50 cases of familial Parkinson’s disease. Can J Neurol Sci 1983;10(1):37–42.

7. Sidransky E, Nalls MA, Aasly JO, et al. Multicenter analysis of glucocerebrosidase mutations in Parkinson’s disease. N Engl J Med 2009;361(17):1651–1661.

8. Cilia R, Tunesi S, Marotta G, et al. Survival and dementia in GBA-associated Parkinson’s disease: The mutation matters. Ann Neurol 2016;80(5):662–673.

9. Liu G, Boot B, Locascio JJ, et al. Specifically neuropathic Gaucher’s mutations accelerate cognitive decline in Parkinson’s. Ann Neurol 2016;80(5):674–685.

10. Anheim M, Elbaz A, Lesage S, et al. Penetrance of Parkinson disease in glucocerebrosidase gene mutation carriers. Neurology 2012;78(6):417–420.

11. Rana HQ, Balwani M, Bier L, Alcalay RN. Age-specific Parkinson disease risk in GBA mutation carriers: information for genetic counseling. Genet Med 2013;15(2):146–149.

12. Balestrino R, Tunesi S, Tesei S, Lopiano L, Zecchinelli AL, Goldwurm S. Penetrance of Glucocerebrosidase (GBA) Mutations in Parkinson’s Disease: A Kin Cohort Study. Mov Disord 2020;35(11):2111–2114.

13. Alcalay RN, Dinur T, Quinn T, et al. Comparison of Parkinson risk in Ashkenazi Jewish patients with Gaucher disease and GBA heterozygotes. JAMA Neurol 2014;71(6):752–757.

14. Driver JA, Logroscino G, Gaziano JM, Kurth T. Incidence and remaining lifetime risk of Parkinson disease in advanced age. Neurology 2009;72(5):432–438.

15. Elbaz A, Bower JH, Maraganore DM, et al. Risk tables for parkinsonism and Parkinson’s disease. J Clin Epidemiol 2002;55(1):25–31.

16. Licher S, Darweesh SKL, Wolters FJ, et al. Lifetime risk of common neurological diseases in the elderly population. J Neurol Neurosurg Psychiatry 2019;90(2):148–156.

17. Gan-Or Z, Liong C, Alcalay RN. GBA-Associated Parkinson’s Disease and Other Synucleinopathies. Curr Neurol Neurosci Rep 2018;18(8):44.

18. Duran R, Mencacci NE, Angeli AV, et al. The glucocerobrosidase E326K variant predisposes to Parkinson’s disease, but does not cause Gaucher’s disease. Mov Disord 2013;28(2):232–236.

19. Mallett V, Ross JP, Alcalay RN, et al. GBA p.T369M substitution in Parkinson disease: Polymorphism or association? A meta-analysis. Neurol Genet 2016;2(5):e104.

20. Lwin A, Orvisky E, Goker-Alpan O, LaMarca ME, Sidransky E. Glucocerebrosidase mutations in subjects with parkinsonism. Mol Genet Metab 2004;81(1):70–73.

21. Cunningham F, Amode MR, Barrell D, et al. Ensembl 2015. Nucleic Acids Res 2015;43(Database issue):D662-669.

22. den Dunnen JT, Dalgleish R, Maglott DR, et al. HGVS Recommendations for the Description of Sequence Variants: 2016 Update. Hum Mutat 2016;37(6):564–569.

23. Thaler A, Bregman N, Gurevich T, et al. Parkinson’s disease phenotype is influenced by the severity of the mutations in the GBA gene. Parkinsonism Relat Disord 2018;55:45–49.

24. Karczewski KJ, Francioli LC, Tiao G, et al. The mutational constraint spectrum quantified from variation in 141,456 humans. Nature 2020;581(7809):434–443.

25. Rentzsch P, Witten D, Cooper GM, Shendure J, Kircher M. CADD: predicting the deleteriousness of variants throughout the human genome. Nucleic Acids Res 2019;47(D1):D886–D894.

26. Davydov EV, Goode DL, Sirota M, Cooper GM, Sidow A, Batzoglou S. Identifying a high fraction of the human genome to be under selective constraint using GERP++. PLoS Comput Biol 2010;6(12):e1001025.

27. Yahalom G, Greenbaum L, Israeli-Korn S, et al. Carriers of both GBA and LRRK2 mutations, compared to carriers of either, in Parkinson’s disease: Risk estimates and genotype-phenotype correlations. Parkinsonism Relat Disord 2019;62:179–184.

28. Chen Y, Gu X, Ou R, et al. Evaluating the Role of SNCA, LRRK2, and GBA in Chinese Patients With Early-Onset Parkinson’s Disease. Mov Disord 2020;35(11):2046–2055.

29. Li N, Wang L, Zhang J, et al. Whole-exome sequencing in early-onset Parkinson’s disease among ethnic Chinese. Neurobiol Aging 2020;90:150 e155–150 e111.

30. Yu Z, Wang T, Xu J, et al. Mutations in the glucocerebrosidase gene are responsible for Chinese patients with Parkinson’s disease. J Hum Genet 2015;60(2):85–90.

31. Balwani M, Fuerstman L, Kornreich R, Edelmann L, Desnick RJ. Type 1 Gaucher disease: significant disease manifestations in “asymptomatic” homozygotes. Arch Intern Med 2010;170(16):1463–1469.

32. Tsuji S, Martin BM, Barranger JA, Stubblefield BK, LaMarca ME, Ginns EI. Genetic heterogeneity in type 1 Gaucher disease: multiple genotypes in Ashkenazic and non-Ashkenazic individuals. Proc Natl Acad Sci U S A 1988;85(7):2349–2352.

33. Huang Y, Deng L, Zhong Y, Yi M. The Association between E326K of GBA and the Risk of Parkinson’s Disease. Parkinsons Dis 2018;2018:1048084.

34. Berge-Seidl V, Pihlstrom L, Maple-Grodem J, et al. The GBA variant E326K is associated with Parkinson’s disease and explains a genome-wide association signal. Neurosci Lett 2017;658:48–52.

35. Wan L, Hsu CM, Tsai CH, Lee CC, Hwu WL, Tsai FJ. Mutation analysis of Gaucher disease patients in Taiwan: high prevalence of the RecNciI and L444P mutations. Blood Cells Mol Dis 2006;36(3):422–425.

36. Goker-Alpan O, Schiffmann R, Park JK, Stubblefield BK, Tayebi N, Sidransky E. Phenotypic continuum in neuronopathic Gaucher disease: an intermediate phenotype between type 2 and type 3. J Pediatr 2003;143(2):273–276.

37. Ida H, Rennert OM, Iwasawa K, Kobayashi M, Eto Y. Clinical and genetic studies of Japanese homozygotes for the Gaucher disease L444P mutation. Hum Genet 1999;105(1-2):120–126.

